# Advancing primary care for childhood pneumonia: a machine learning-based approach to prognosis and case management

**DOI:** 10.1101/2024.02.22.24303209

**Authors:** Oguzhan Serin, Izzet Turkalp Akbasli, Sena Bocutcu Cetin, Busra Koseoglu, Ahmet Fatih Deveci, Muhsin Zahid Ugur, Yasemin Ozsurekci

**Author notes:** **Corresponding author:** Izzet Turkalp AKBASLI, MD, Department of Pediatrics, Hacettepe University Medical School, Ankara, 06230, Türkiye, **E-mail:****, **.

## Abstract

**Background:** Pneumonia is the leading cause of preventable mortality under five years of age. Appropriate case management is as essential as disease prevention interventions, especially in primary care settings. Computer science has been used accurately and widely for pneumonia diagnosis; however, prognosis studies are relatively low. Herein, we developed a machine learning-based clinical decision support system tool for childhood pneumonia to provide prognostic support for case management.

**Methods:** We analyzed data from 437 children admitted to our clinic with a pneumonia diagnosis between 2014 and 2020. Pediatricians encoded the raw dataset according to candidate features. Before the experimental study of the machine learning algorithms of Pycaret, SMOTE-Tomek was utilized for managing imbalanced datasets. The feature selection was made by examining the SHAP values of the algorithm with the highest performance and re-modeled with the most important clinical features. We optimized hyperparameters and employed ensemble methods to develop a robust predictive model.

**Results:** Optimized models predicted pneumonia prognosis with %77-88 accuracy. It was shown that severity could be determined over %84 by five clinical features: hypoxia, respiratory distress, age, Z score of weight for age, and antibiotic usage before admission.

**Conclusions:** In this experimental study, we demonstrated that contemporary data science methods, such as oversampling, feature selection, and machine learning tools, are promising in predicting the critical care need of patients. Even in small-size samples like our study, ML methods can reach current wisdom.

**Highlights:**

- Pneumonia accounts for 14% of mortality in children under 5, with over 740,000 deaths in 2019 alone.
- WHO and UNICEF’s GAPPD aims to cut mortality rates by focusing on vaccinations, sanitation, breastfeeding, and addressing pediatric HIV.
- Accurate diagnosis and timely treatment can reduce pneumonia mortality by up to 28%, yet diagnosing can be challenging.
- Many in underdeveloped regions lack access to essential equipment and trained staff, exacerbating mortality rates.
- Data science and machine learning offer promising solutions for pneumonia management, especially in LMICs, with a focus on prognostic support.

## 1. Background

Pneumonia is responsible for 14% of all mortality under the age of 5 and is included in WHO reports as the cause of death in 740,180 children in 2019 alone [1]. Global Action Plan for the Prevention and Control of Pneumonia and Diarrhea (GAPPD), which was released by WHO and UNICEF, and have aimed to reduce the mortality rate from pneumonia and diarrhea under five years old [2], [3]. They have set targets as vaccination, water, and air sanitation, exclusively breastfeeding in the first six months, and eliminating pediatric HIV cases, along with appropriate pneumonia and diarrhea case management.

It has been demonstrated that timely and accurate diagnosis of pneumonia and appropriately initiated treatment reduces mortality by up to 28% [4], [5]. Diagnosis can often be difficult, since the clinical presentation of pneumonia in children is variable [6]. For this reason, WHO has published the Integrated Management of Pediatrics (IMCI) guidelines, which guide physicians for diagnosis, treatment, danger signs of pneumonia [7]. Childhood pneumonia can easily be preventable and treatable with low-cost and low-tech medication at primary care [1]. However, this preventable health problem continues to be the most important cause of mortality, especially in underdeveloped countries and regions, due to the lack of equipment and trained human resources. In addition, it has been shown that the seeking for health services by families living in these regions causes delays in providing appropriate treatment and causes progression in disease severity [8]. This shows that case management should be improved as well as disease prevention interventions, especially in regions with limited resources.

Data Science can provide actionable evidence for effective clinical intervention in pediatric diseases in the future [9] and can reduce inequality in healthcare [10]. Also, using big data and machine learning technologies is promising for childhood pneumonia case management, especially in low-income and middle-income countries (LMICs) [11]. Because of their flexibility and high accuracy, machine-learning models are used in medicine in the fields of prediction (prognostics) and classification (diagnostics) [9]. It has been seen that the vast majority of data science studies on pneumonia aim to provide diagnostic support to the physician by processing radiologic images [12]. However, diagnostic equipment is mostly unavailable in LMICs and primary care units. Therefore, physicians need prognostic support algorithms that distinguish between serious and non-serious cases.

We aimed to develop machine learning-based clinical decision support system tool for childhood pneumonia that can be used by physicians, particularly working in LMICs in order to ensure the effective case management of pneumonia, which is one of the 2025 goals of WHO [2].

## 2. Material and Methods

### 2.1 Case definition and patient selection

Our study includes pediatric patients who received inpatient treatment at our clinic between January 2014 and April 2020, with the diagnosis of community acquired pneumonia (CAP) according to IMCI guidelines and its latest revision [7], [13]. Patients who were neonatal age, older than 18, and hospitalized within the last 14 days were excluded. This study design and procedures were approved by Hacettepe University Clinical Research Ethics Committee with protocol number GO-20/1182.

The medical records of 437 patients were retrospectively examined by the pediatricians, and they were asked to code according to the candidate features (Table 1). These variables were chosen based on their clinical value in clinical decision-making and their availability in primary care.

**Table 1.** Candidate Features.

| Clinical Variables | Laboratory Variables |
| --- | --- |
| 1. Age | 1. Hemoglobine |
| 2. Weight | 2. Leucocytes |
| 3. Z score | 3. Lymphocytes |
| 4. Gender | 4. Neutrophiles |
| 5. Complaint period | 5. Platelets |
| 6. Comorbidity | 6. C-reactive Protein |
| 7. Recent Antibiotics usage | 7. Albumine |
| 8. Fever | 8. Sodium |
| 9. Cough | 9. Aspartate aminotransferase(AST) |
| 10. Loss of apetit | 10. Alanin aminotransferase(ALT) |
| 11. Respiratory distress |  |
| 12. Abnormal lung sounds |  |
| 13. Hypoxia |  |

The primary outcome was CAP prognosis which is scaled into two-level severe or non-severe. This categorization was made by physician-encoders according to whether the patient required tertiary care referral or not. Children with poor prognoses included those admitted to the pediatric intensive care unit (PICU) and/or required any oxygenation and ventilation support.

### 2.2 Data preprocessing

Data preprocessing, analysis, visualization, and experimental setup of models were developed by using Python 3.9 programming language. We used Python’s libraries Pandas, NumPy, Matplotlib, Seaborn, and Plotly for exploratory data analysis. We employed the PyCaret library for model development. The library is equipped with various preprocessing modules that enable the iterative handling of missing data using the LightGBM algorithm. Furthermore, we identified anomalous data points through the implementation of PyCaret’s unsupervised anomaly detection module. We made use of Pycaret’s classifier module to perform data preprocessing, where we normalized the numerical data using the Min-Max scaler method and applied the one hot encoder function to handle categorical data.

### 2.3 Handling with the imbalanced dataset

When measured balance with the Shannon entropy of the dataset, it resulted in 0.7, this result can be accepted as the imbalanced dataset. We handled the imbalanced dataset with SMOTE-Tomek, a variant of the noteworthy oversampling method named Synthetic Minority Oversampling Technique (SMOTE). This method oversamples the minority class, and Tomek Links removes samples from the majority class with overlapping values. So, the ratio of samples becomes 1:1. We used the Imblearn library for implementing data oversampling.

We split our datasets into two sets using the “train_test_split” method of the SciKit-Learn library. In the beginning, we divided 5% of the general dataset as test data in order to prevent data leakage. The other part of the data (85%) was used for training and validation, 85% and 15%, respectively.

### 2.4 Algorithms

Pycaret provides efficient implementations of state-of-the-art algorithms and is reusable among scientific disciplines. We used the Pycaret Classifier module for classification, which includes the following models: Ridge Classifier (Ridge), Linear Discriminant Analysis (LDA), Naïve Bayes (NB), Extra Tree Classifier (ET), Extreme Gradient Boosting (XGBoost), Random Forest (RF), Gradient Boosting Classifier (GBC), Light Gradient Boosting Machine (LightGBM), CatBoost Classifier (CatBoost), Logistic Regression (LR), K Neighbours Classifier (KNN), Decision Tree (DT), Ada Boost Classifier (AdaBoost), Quadratic Discriminant Analysis (QDA), Support Vector Machine Linear Kernel Classifier (SVM), and Dummy Classifier (Dummy).

In our work, we considered 10-fold cross-validation. While developing our model with Pycaret tools, we implemented the tuning function using the Tune-Sklearn library and the ‘hyper-band’ optimization algorithm to obtain a set of best-performing parameters. For ensembling, we also used Pycaret classifier ensemble, stack, and blender methods. Ensembling methods have strong evidence that they can significantly enhance the accuracy of classifications. After the optimization of parameters, in the last phase, we utilized the most common ensemble methods provided by the Pycaret library to further improve our model’s performance (Figure-1).

**Figure-1:**
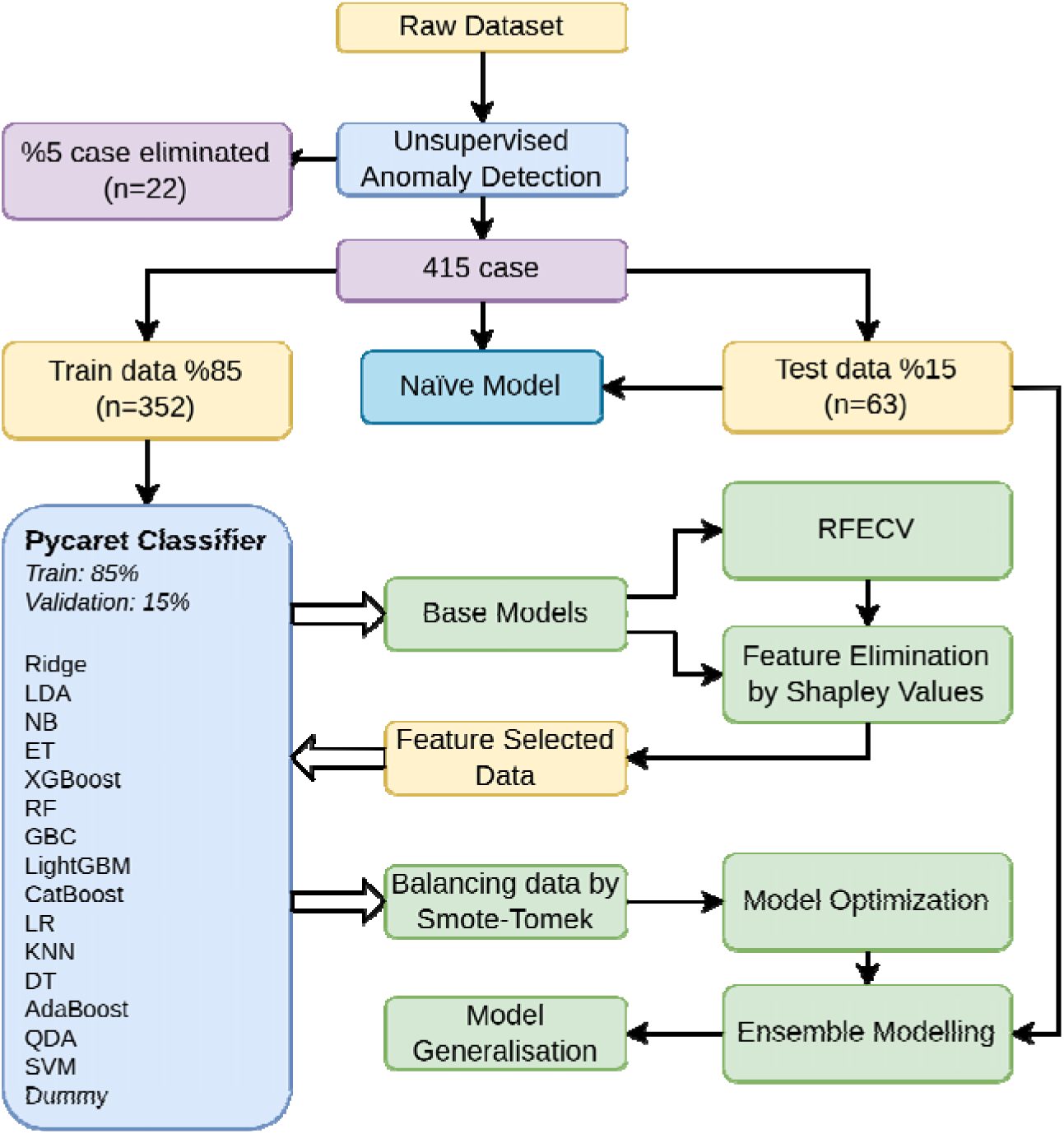
The Experimental Setup : In this figure, we illustrate the experimental process of our models. Initially, we cleaned the data by identifying 5% of cases as abnormal data using unsupervised learning. We then split the data into a Train set (85%) and a Validation set (15%) using the Pycaret classifier model. The base model with the highest ROC-AUC value was the RandomForest algorithm. Subsequently, we determined the optimal number of features as 18 using RFECV and selected the top 18 features based on Shapley values. We then balanced the dataset using the SMOTE-Tomek method and developed high-performing models. After optimizing the hyperparameters, we selected the best-performing model and created new models by using ensemble methods. In parallel, we developed a new model using only clinical findings for clinical prediction. For abbreviations, Ridge Classifier (Ridge), Linear Discriminant Analysis (LDA), Naïve Bayes (NB), Extra Tree Classifier (ET), Extreme Gradient Boosting (XGBoost), Random Forest (RF), Gradient Boosting Classifier (GBC), Light Gradient Boosting Machine (LightGBM), CatBoost Classifier (CatBoost), Logistic Regression (LR), K Neighbours Classifier (KNN), Decision Tree (DT), Ada Boost Classifier (AdaBoost), Quadratic Discriminant Analysis (QDA), Support Vector Machine Linear Kernel Classifier (SVM), Dummy Classifier (Dummy).

### 2.5 Feature Selection and data-reducing methods

Feature selection is a process of one-by-one evaluation to determine which features are effective on the result within the dataset. Irrelevant or partially relevant features can negatively impact ML model performance and make the ML model learn based on irrelevant features. These methods are aimed at eliminating irrelevant features and keeping the strong features to reduce the dimension of the dataset. Recursive feature elimination (RFE) is a feature selection method that fits a model and removes the irrelevant features until the specified number of features is reached. Recursive feature elimination with cross-validation (RFECV) aims to select the optimal number of features with permutation importance and recursive feature elimination. In this study, we used the RFECV module from “yellowbrick” library for selecting the optimum feature number. SHAP (SHapley Additive exPlanations) methodology is an innovative tool for explaining machine learning decision-making processes for datasets. The goal of the SHAP method is to present and explain the prediction with respect to the contribution of each feature to the predicted value. In RFECV, the features are ranked by a permutation importance measure. The SHAP algorithm’s importance value is able to enforce consistency and accuracy more than the permutation approach, so we have used SHAP library algorithms for feature selection (Figure 2). Ultimately, RFECV algorithms showed us 18 parameters are sufficient to explain nearly 90% of variances. A total number of 13 clinical and 5 laboratory variables were selected according to their SHAP values (Figure 2).

**Figure-2:**
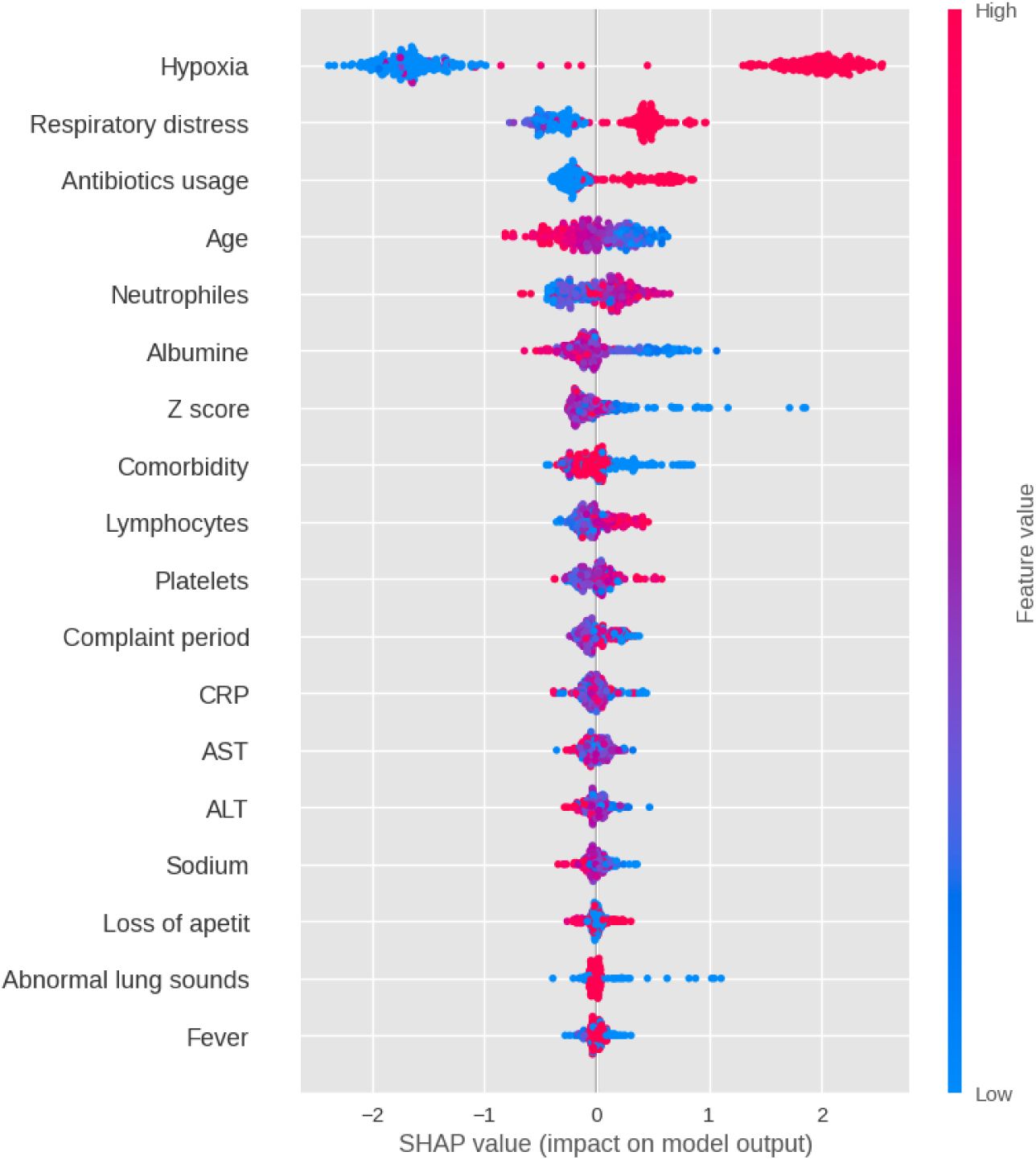
Feature selection Algorithms: In Figure 2, we present the results of SHAP values which is for the RandomForest Classifier model with the highest ROC-AUC score in the dataset before feature selection, using the SHAP library’s plot_summary module. The y-axis shows the importance of each feature, with the most important feature at the top and the least important at the bottom. The colors may represent the contribution of each feature to the model’s prediction. For example, features that have a large positive contribution to the prediction may be shown in a warm color (e.g. red), while features that have a large negative contribution may be shown in a cool color (e.g. blue). In this example, hypoxia is the most important attribute in the plot. The presence of hypoxia (hypoxia = 1) causes the model to move closer to the target class, while its absence causes the model to move away from the target class. This predicts that hypoxia is an aggravating factor. On the other hand, evaluating the albumin attribute shows that high levels of albumin are protective for the target class. In summary, hypoxia is an adverse factor, and high albumin levels are protective.

## 3. Results

In this section, we present a comparison of the performance of 16 different algorithms for raw and pre-processed datasets. We used various evaluation metrics such as accuracy, area under the receiver operator characteristic curve (AUC), recall, precision, F1 score, Cohen’s kappa (Kappa), and MCC (Matthews correlation coefficient) to assess model performance. To analyze model performance, all prediction experiments were conducted using ten-fold cross-validation. Subsequently, the models were optimized, and their performances were evaluated on a balanced dataset using SMOTE-Tomek and feature selection. The performances of the three models with the highest performance (CatBoost, XGBoost, and LightGBM) were evaluated by applying hyperparameter optimization and ensemble methods. Table-2 compares the results obtained with CatBoost, XGBoost, and LightGBM among the optimized and non-optimized results and the results of the combinations with the highest performance from the basic ensembling methods (ensembling, blending, and stacking methods). The highest AUC value was achieved by using Optimized LightGBM as the meta-model in the Stacking method.

**Table-2 :**
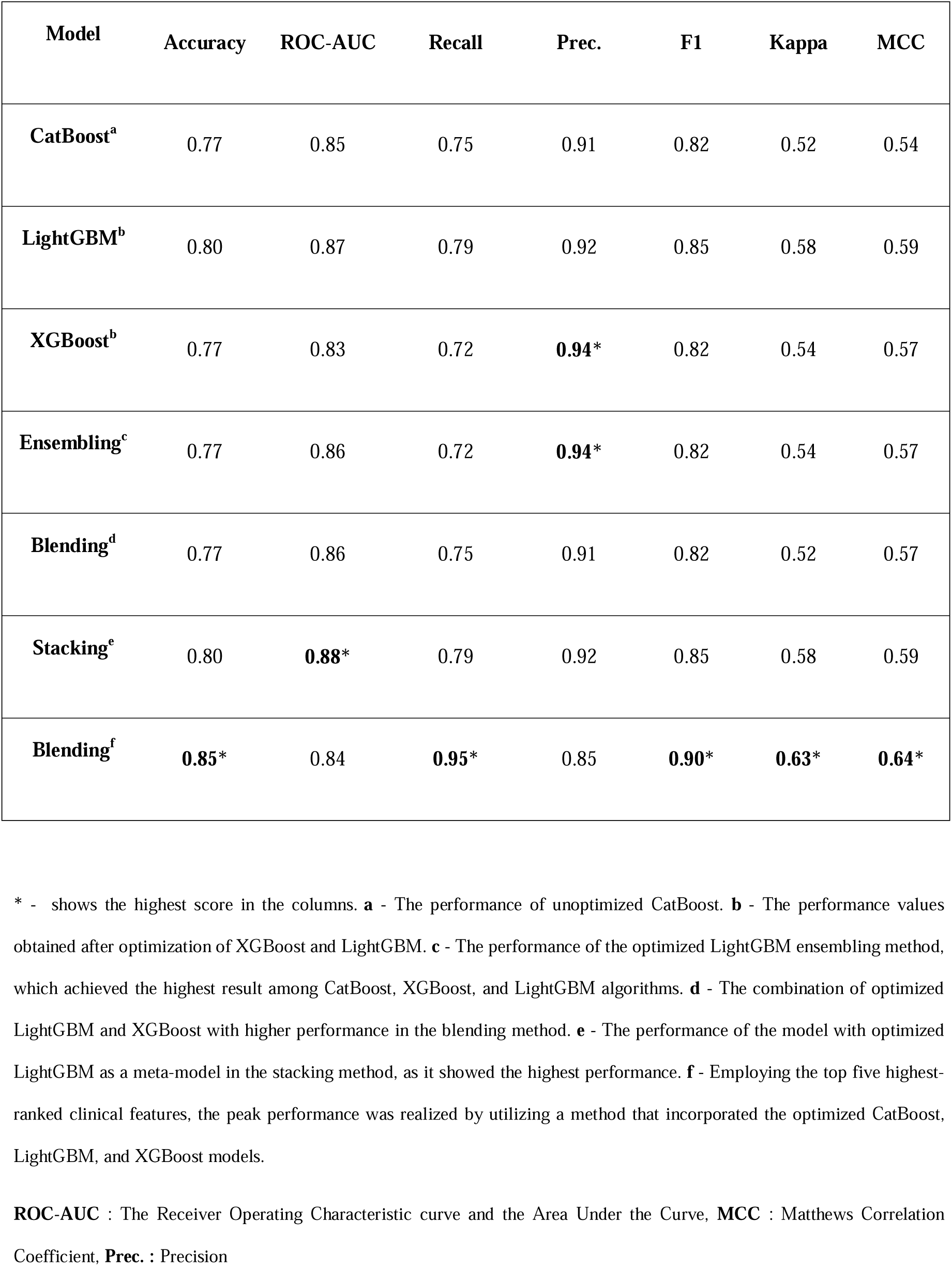
Model Performance Results.

The optimized LightGBM in the model, developed with balanced and feature-selected data, was responsible for the attainment of the highest performance. Upon evaluation of clinical features according to SHAP values, a ranking was established based on their feature importance scores, with the highest score being garnered by the top five clinical features (hypoxia, respiratory distress, age, Z score of weight for age, and antibiotic usage before admission) (Supplementary - 1). The application of a workflow employing these five features, as done previously, resulted in the highest performance (84%), which was achieved through the utilization of the ensemble method, incorporating the blending method of the optimized CatBoost, LightGBM, and XGBoost models.

## 4. Discussion

Pneumonia, the leading cause of childhood mortality, is also one of the most common causes of hospitalization [3], [14]. Herein we present a contemporary approach to building an ML-based case management support tool that assists primary care physicians in determining where the case should be managed with an accuracy of more than 80%.

The leading causes of this disease are still viral infections and co-infections in both children/adults and high/low-income countries [15]. In most cases, transmission occurs through close contact and inhalation of infected droplets [1]. Since March 2020, COVID-19 outbreak measures have indirectly decreased the incidence of non-COVID pneumonia and other respiratory-borne diseases by preventing transmission [16], [17].

Today, there is a wide knowledge of the prevention, diagnosis, treatment, and management of complications in childhood pneumonia, but due to resource limitations, it is not possible for all physicians and patients to benefit from this. During the worldwide pandemic, data-driven solutions to global concerns were implemented [18]. This provides us with a different viewpoint on non-covid pneumonia case management.

Since March 2020, the declaration of a global pandemic by the World Health Organization, a substantial amount of data about COVID-19 has been published [19]. There have been numerous COVID-related AI studies focused on pneumonia diagnosis by radiological findings [20]. However, pneumonia diagnosis is clinical, and routine chest radiographs are not necessary for the confirmation diagnosis [21]and do not improve outcomes [22]. In addition, chest radiography (CXR) can be used only in inpatient settings to identify complications or evaluate response to treatment.

AI studies aiming to create a decision support system are often on either risk prediction (prognostics) or classification (diagnostics) [9]. Although strong diagnostic support algorithms have been published in pneumonia-related studies in recent years, there is still a need for prognosis prediction studies for case management [20]. Determining the severity of a disease or predicting its prognosis answers essential questions of physicians in medical decision-making, such as “*Where should it be treated? Outpatient? ICU?*”, “*Which therapy should I start? How long should I give it?*”, “*When should I discharge the patient? When should I call for control?*”. There are several studies and guidelines in the literature for severity assessment and prognosis prediction of pneumonia [23]. For the majority, mortality and/or development of complications were the primary outcomes, and clinical, radiological, and laboratory variables are the key predictors.

This study reviewed important clinical data and prognostic outcomes of pneumonia patients hospitalized in a major tertiary hospital before the pandemic. Two pediatricians from our research team were only in charge of labeling and encoding the data from the records of hospitalized children. Other pediatricians were involved in data analysis, model development, and literature review. Our main outcome was to assess the necessity of referral to tertiary care. For this reason, we asked physicians to encode clinical and laboratory data that can be obtained in primary care settings.

In this study, we balanced the dataset with SMOTE-Tomek and, selected features by SHAP values, then conducted experiments with a variety of advanced machine learning algorithms on this new structured dataset. Since our raw dataset was imbalanced in terms of outcome variables.

Imbalanced datasets can reduce the ML algorithm’s performance. For example, if models are prepared with imbalanced data, results will be more likely in the majority class. The minority class of target has a low precision value, which led to an accuracy paradox, as previously shown [24]. To overcome this issue, re-sampling methods have already been widely implemented in order to reduce the weight of the majority class to the minority class. However, there is a possibility of losing valuable medical data in this way. One of the proposed re-sampling methods is SMOTE. Recently, there are studies that support SMOTE-Tomek, a variant method of SMOTE, which can be effectively employed in preprocessing imbalanced medical data for the prediction of disease classification [25], [26].

In terms of feature selection, there may be certain variables in the dataset that are not beneficial for the model-developing process. These low-importance features create noise and reduce the precision of the model. Therefore, we determined the optimum number of features with RFECV to minimize noise. RFECV was already employed in medical research, and it was successful in our study as well [27].

Conventional feature selection methods include a variety of post-hoc analyses. New feature selection techniques have been introduced in conjunction with the development of ML algorithms. We used SHAP values to explain the ML model’s results in part because they are more robust for categorical and numerical variables and more commonly utilized in clinical data science. Furthermore, articles have been published promoting how SHAP is superior to the others, such as Local Interpretable Model-agnostic Explanations (LIME) [28]–[30].

The main outcome of this study was not only to create the best model but also to answer the question for a primary care physician *“Where the case should be managed in?”*. We achieved a score of 84% (Supp. Figure 1.) with the five most important clinical findings such as hypoxia, respiratory distress, age, fever and complaint period. It was be observed that these variables are already involved in previous prognosis studies [22], [23], [31], [32]. It could be a life-saving decision for an inexperienced doctor, who uses these five features in limited resources, to foresee the critical care need of the patient.

In conclusion, we applied contemporary data pre-processing and ML methods to our dataset and developed substantially successful models. While investigating feature importance that enhances the performances of models, we encountered the same variables which are already considered to be prognosis predictors by previous studies and guidelines. Even in small-size samples like in our study, ML methods can reach current wisdom. Since ML algorithms facilitate the transformation and purification of expert knowledge for universal applications, we think also have a great deal of potential for other diseases.

## Data Availability

All data produced in the present study are available upon reasonable request to the authors

## Author contributions

**Oguzhan Serin, Pediatrician, M.D. :** Creation of the work plan, interpretation of statistical analysis and machine learning algorithm, co-investigate the literature, writer of the background and discussion

**Izzet Turkalp Akbasli, Pediatrician, M.D. :** Building machine learning algorithms and co-investigate the literature, writer of the results and methods.

**Sena Bocutcu, Pediatrician, M.D. :** Scanning patients from the Hospital Electronic Health Record System, encoding the attributes of the patients’ data in the case report form. “Human Encoder-1”

**Busra Koseoglu, Pediatrician, M.D. :** Scanning patients from the Hospital Electronic Health Record System, encoding the attributes of the patients’ data in the case report form. “Human Encoder-2”

**Ahmet Fatih Deveci, M.S. Engineer:** Building machine learning algorithms, Optimizing the dataset.

**Muhsin Zahid U**Ğ**UR, PhD :** Coding of advanced statistical and machine learning algorithms, creation of clinical decision support system interface.

**Yasemin Ozsurekci, Pediatrician, M.D. :** Creation of the work plan, interpretation of statistical analysis, gathering the team of investigators.

## Acknowledge

None

## Further considerations

The manuscript has been thoroughly ‘spell-checked’ and ‘grammar-checked’. All references listed in the Reference List are duly cited within the text, and conversely, every citation in the text has a corresponding entry in the Reference List. The authors have reviewed and adhered to the journal policies as detailed in the provided guide.

## Summary Table

| What was already known on this topic | What this study added to our knowledge |
| --- | --- |
| Many AI medical studies look at radiological images, but pneumonia is mainly a clinical diagnosis. Also, primary care often lacks this imaging equipment. | In our study, using clinical characteristics, an AI algorithm predicts patient prognoses with 85% accuracy, emphasizing the need for prognostic support in regions lacking advanced diagnostic tools. |
| In conventional data analyses, if data imbalance is not balanced, prediction models are biased towards the majority group. | In this study, the SMOTE method was used to balance the data, which in turn enhanced the model's generalizability. |
| Many valuable studies are either confined to their specific centers or the tools developed are not made accessible to other healthcare professionals. | The models in this study have been deployed online and can be accessed at "https://pnm.pediatridefteri.com", and this represents a potential publishing initiative. |

## Declaration of Generative AI and AI-assisted technologies in the writing process

During the preparation of this work, the authors utilized OpenAI GPT-4 to restructure sentences for enhanced readability, as they are not native English speakers. After using this tool/service, the authors reviewed and edited the content as needed and took full responsibility for the content of the publication.

