## Supplementary - 1 for "Advancing primary care for childhood pneumonia: a machine learning-based approach to prognosis and case management"

- **Supplementary Figure 1**: SHAP values forward selection method
- **Supplementary Table 1**: Statistical inferences of candidate variable between severe and non-severe pneumonia case groups

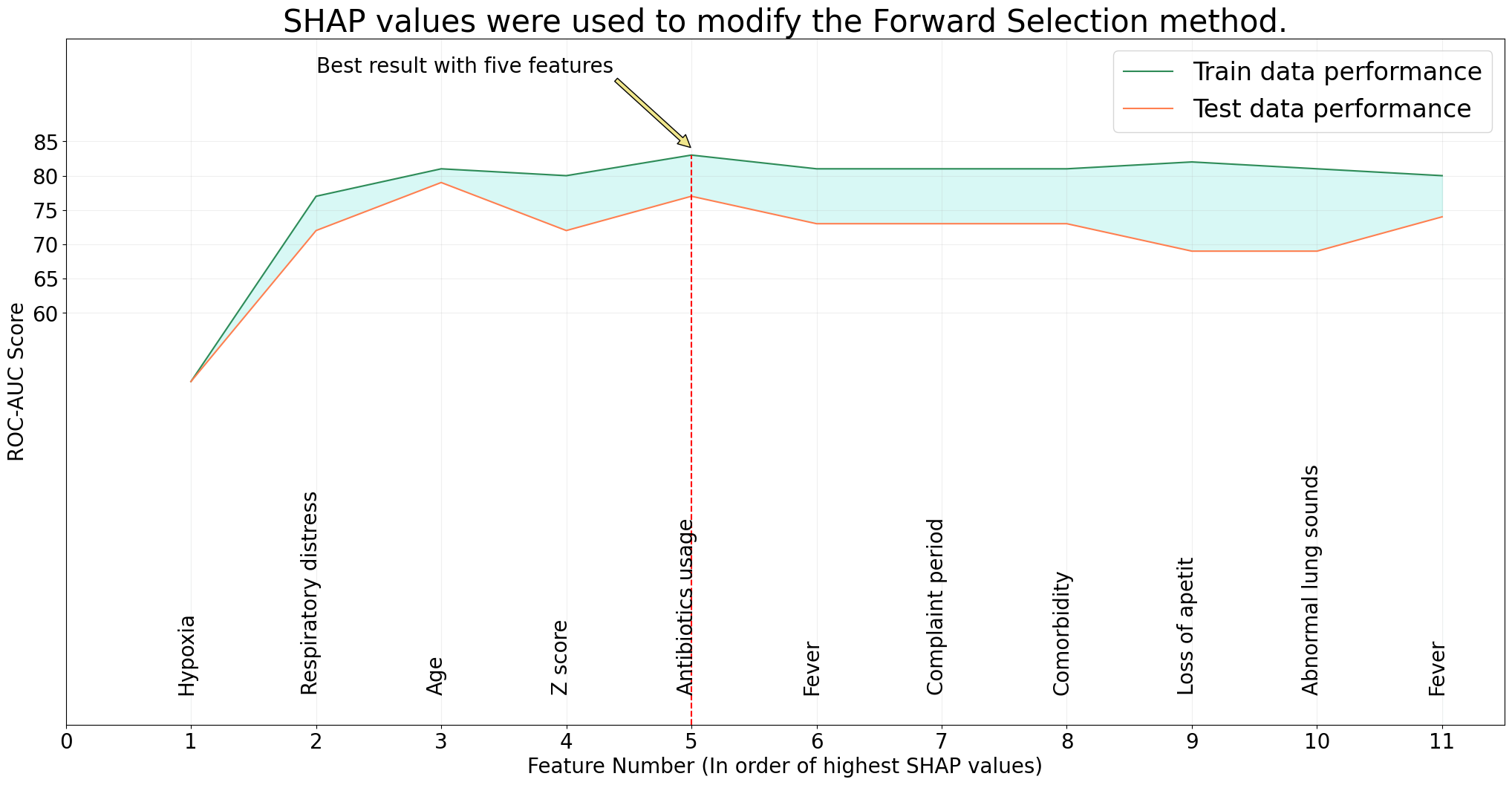

**Supplementary Figure 1**: SHAP values forward selection method

| **Candidate Variables** | **Non-severe**  **(n=133, %30,4)** | **Severe**  **(n=304,%69,6)** | **Statistic^a,b^** | **Significance** |
| --- | --- | --- | --- | --- |
| **Age (months)** | 44 (13-98) | 23 (7-64,5) | 16602^b^ | 0,003* |
| **Weight (Z scores)** | -0,57(-1,4-0,45) | -0,7(-2,5-0,4) | 17784^b^ | 0,045* |
| **Complaint period (days)** | 4(2-7) | 4(2-7) | 19274 ^b^ | 0,435 |
| **Gender** |  |  |  |  |
| Male | 68(%30,9) | 152(%69,1) |  |  |
| Female | 65(%30) | 152(%70) | 0,047^a^ | 0,828 |
| **Comorbidity** |  |  |  |  |
| No | 48(%34) | 93(%66) |  |  |
| Yes | 85(%28,7) | 211(%71,3) | 1,280^a^ | 0,258 |
| **Recent Antibiotic Usage** |  |  |  |  |
| No | 93(%32,6) | 192(%67,4) |  |  |
| Yes | 40(%26,3) | 112(%73,7) | 1,868^a^ | 0,172 |
| **Fever** |  |  |  |  |
| No | 33(%26) | 94(%74) |  |  |
| Yes | 100(%32,3) | 210(%67,7) | 1,675^a^ | 0,196 |
| **Cough** |  |  |  |  |
| No | 18(%26,1) | 51(%73,9) |  |  |
| Yes | 115(%31,3) | 253(%68,8) | 0,508^a^ | 0,476 |
| **Loss of appetit** |  |  |  |  |
| No | 96(%30) | 224(%70) |  |  |
| Yes | 37(%31,6) | 80(%68,4) | 0,107^a^ | 0,744 |
| **Respiratory Distress** |  |  |  |  |
| No | 90(%48,4) | 96(%51,6) |  |  |
| Yes | 43(%17,1) | 208(%82,9) | 49,295^a^ | <0,001* |
| **Abnormal Lung Sounds** |  |  |  |  |
| No | 31(%53,4) | 27(%46,6) |  |  |
| Yes | 102(%26,9) | 277(%73,1) | 16,729^a^ | <0,001* |
| **Hypoxia** |  |  |  |  |
| No | 113(%63,8) | 64(%36,2) |  |  |
| Yes | 20(%7,7) | 240(%92,3) | 156,818^a^ | <0,001* |
| **Hemoglobine (gr/dl)** | 11,6(10,4-12,9) | 11,6(10,6-12,6) | 20022,5 ^b^ | 0,873 |
| **Leukocytes (x10^6/L)** | 9900(6800-14600) | 10950(8050-15850) | 17837 ^b^ | 0,05* |
| **Lymphocytes (x10^6/L)** | 2300(1400-3700) | 2800(1900-4400) | 17039 ^b^ | 0,01* |
| **Neutrophiles (x10^6/L)** | 5285(2700-9200) | 6500(3650-10900) | 17645,5 ^b^ | 0,045* |
| **Platelets (x10^9/L)** | 310(225-386) | 317,5(230,5-425) | 19399 ^b^ | 0,501 |
| **C-Reactive Protein (mg/L)** | 2,06(0,79-7,67) | 2,06(0,83-7,35) | 19842,5 ^b^ | 0,758 |
| **Albumin (gr/dl)** | 3,9(3,73-4,2) | 3,9(3,4-4,2) | 17121 ^b^ | 0,011* |
| **Sodium(meq/L)** | 136(135-138) | 136(134-138) | 19657,5 ^b^ | 0,644 |
| **Aspartat transaminase (U/L)** | 35(26-42) | 35(28-50) | 18382 ^b^ | 0,131 |
| **Alanine transaminase (U/L)** | 17(12-26) | 18(13-29) | 18457 ^b^ | 0,147 |
| “*” shows the significant differences between two groups.  a: Chi-square value b: Mann-Whitney U test  Numerical variables were showed in median (interquartile range) | | | | |

**Supplementary Table 1**: Statistical inferences of candidate variable between severe and non-severe pneumonia case groups
